# Pan genome clustering identifies a novel mosaic prophage specific to *Salmonella* Enteritidis lineage associated with the invasive disease in India

**DOI:** 10.1101/2025.05.08.25327125

**Authors:** Jobin John Jacob, Aravind Velmurugan, Dhanalakshmi Solaimalai, Ramya Iyadurai, Karthik Gunasekaran, Jansi Rani Malaiyappan, M Yesudoss, M Vasuki, Binesh Lal, Jacob John, Kamini Walia, Balaji Veeraraghavan

**Author notes:** Corresponding author Dr Balaji Veeraraghavan Professor Department of Clinical Microbiology, Christian Medical College, Vellore, Tamil Nadu, India, 632004. These authors contributed equally to this work (Jobin John Jacob, Aravind Velmurugan).

## Abstract

*Salmonella enterica* serovar Enteritidis is a leading cause of bloodstream infections (BSIs), particularly in sub-Saharan Africa (sSA). While typhoidal salmonellosis is well documented in South Asia, the burden and genomic characteristics of non-typhoidal *Salmonella* (NTS)-associated BSIs remain understudied. This study investigates the clinical presentation, phylogenetic relationships, invasive potential, and transmission dynamics of *S.* Enteritidis causing BSIs in India. Clinical data were collected from 101 bloodstream infection cases caused by *S*. Enteritidis (2012–2022). Representative isolates of *S*. Enteritidis originating from other clinical samples and livestock samples (*n=17*) were also included for comparative analysis. Study isolates were subjected to whole genome sequencing (WGS) using the Illumina platform and phylogenetic structure, invasiveness index, prophage profiles, and transmission dynamics were analyzed through comparative genomics. Clinical data indicated that infants and immunosuppressed individuals were at the highest risk of *S.* Enteritidis BSI. Phylogenetic analysis revealed four major lineages, with most study BSI isolates clustering in the Global Intermediate Clade, a novel lineage defined by our study.. This clade exhibited the second-highest invasiveness index (median: 0.221, SD: 0.013), following the West African Clade (median: 0.253, SD: 0.25). Isolates originated from poultry samples clustered separately and were not associated with human BSI strains. Pan-genome analysis revealed that the Global Intermediate Clade acquired a novel mosaic prophage, incorporating genetic elements from *Salmonella* and *Vibrio* phages, suggesting a role in host adaptation and virulence. This study provides contemporary insights into the genomic and clinical dynamics of *S.* Enteritidis BSIs in India. The distinct phylogenetic placement, high invasiveness index, and unique prophage acquisition of the Global Intermediate Clade indicate its potential for extraintestinal adaptation and human-specific transmission. Further research is required to delineate environmental reservoirs and transmission pathways contributing to the persistence of this emerging lineage.

**Author Summary:** In this study, we investigated *Salmonella enterica* serovar Enteritidis, a major cause of invasive bloodstream infections (BSIs), particularly among infants and immunocompromised individuals in India. While invasive non-typhoidal Salmonella (iNTS) is well-documented in sub-Saharan Africa, its burden in South Asia remains understudied. By analyzing clinical and genomic data from BSI patients over a decade, we identified a distinct genetic lineage associated with BSI infections, which we named the “Global Intermediate Clade.” This lineage harbors a unique mosaic prophage—a viral DNA segment integrated into its genome—that may enhance its ability to cause invasive disease. Our findings show that this clade has a high potential for causing severe infections, similar to strains found in Africa. Notably, *S*. Enteritidis isolates from poultry in India were genetically distinct from those causing BSI, suggesting limited direct transmission from animals to humans. This highlights the need for enhanced genomic surveillance to track emerging pathogen lineages and understand their evolution. By identifying this previously unrecognized clade, our study provides critical insights that could inform public health strategies for controlling Salmonella infections in high-risk populations. Future research should explore the role of this prophage in virulence and its implications for vaccine and treatment development.

## Introduction

Nontyphoidal *Salmonella* (NTS) are a significant cause of gastrointestinal infections which are generally self-limiting and resolve without any clinical interventions [1, 2]. However, over the past few decades, some serovars of NTS have emerged as a leading cause of BSI particularly in sub-Saharan African (sSA) countries [3, 4]. As of 2017, sub-Saharan Africa accounted for 79% of the 535,000 global cases of invasive NTS (iNTS) disease and 85% of the 77,500 associated deaths [5]. iNTS infections are strongly associated with immunocompromised individuals, including those with HIV, malaria, malnutrition, and hematologic malignancies [6, 7]. Clinical presentation often includes non-specific febrile illness, closely resembling typhoid fever or malaria, making diagnosis particularly challenging in resource-limited settings [8].

Although a wide range of *Salmonella* serovars have been reported to cause iNTS diseases, the majority of infections in sub-Saharan Africa are caused by two dominant serovars: *Salmonella enterica* serovar Typhimurium (ST313) and *Salmonella enterica* serovar Enteritidis (ST11) [9,10]. These endemic African lineages have been identified as the primary drivers of iNTS infections in the region [11]. Phylogenetic analysis based on single nucleotide polymorphisms (SNPs) has identified three distinct lineages (L1, L2, and L3) of *S*. Typhimurium ST313, which have evolved over time to exhibit increased human adaptation and invasiveness [12]. Similarly, the African clade of ST11 *S*. Enteritidis has been predicted to be more invasive than the global epidemic clone and global outlier cluster isolates [13].

While iNTS disease in sub-Saharan Africa has been studied extensively, isolates from other low-income settings have received less attention. In South Asian countries, the prevalence of iNTS has been largely underestimated due to limited surveillance data and underreporting [14, 15]. Furthermore, the transmission dynamics of NTS serovars between humans, animals, and the environment in a One Health context remain poorly characterized [16]. Previously, a retrospective analysis of blood cultures collected at our tertiary care hospital in southern India region between 2000 and 2020 revealed that *S*. Typhimurium (49%) and *S*. Enteritidis (29%) were the most prevalent serovars causing iNTS infections [17]. Of these two, *S*. Typhimurium has been relatively well characterized, largely due to its use as an experimental model for human typhoid fever. However, the clinical presentation, antimicrobial resistance patterns, and genomic features of *S*. Enteritidis infections in our setting remain poorly documented.

In this study, we conducted a retrospective analysis of *S*. Enteritidis isolates obtained from BSI at a tertiary-care hospital in Vellore, India. Clinical characteristics and disease outcomes of these infections were also assessed. Additionally, *S*. Enteritidis isolates from livestock sources were included to investigate potential zoonotic transmission dynamics between humans and animals. Whole-genome sequencing (WGS) and comparative genomic analysis were performed to determine the phylogenetic relationships, genetic diversity, and invasive potential of these isolates within a global isolate collection. Furthermore, pan-genome analysis was performed to shed to understand evolutionary trajectory, host adaptation mechanisms, and pathogenicity determinants of *S*. Enteritidis, in order to shed light on its potential for human infection and dissemination.

## Methods

### Study design and data collection

This retrospective study was conducted between January 2012 and December 2022 at the Christian Medical College (CMC), Vellore, India. CMC Vellore is a 2,234-bed, multispecialty, tertiary-care teaching hospital that serves as a referral center for various medical disciplines. Data were retrieved from the hospital’s electronic medical records system (Clinical Workstation). Clinical information was collected for patients admitted with *Salmonella* Enteritidis bloodstream infections (BSI), including sociodemographic details, presenting clinical features, co-infections, antibiotic use, and patient outcomes. Additionally, underlying conditions such as HIV status, autoimmune disorders, chronic diseases, malignancies, and the use of steroids, cytotoxic drugs, and other relevant comorbidities were documented. Patient outcomes including length of hospital stay, discharge status, and mortality rates were also assessed. All the patient identifying variables in the data were removed before the analysis. This study received ethical approval from the Institutional Research Board and Ethics Committee of CMC Vellore (IRB Min. No. 11878 dated 27^th^ February, 2019). As the required data had been collected as part of the standard of care for diagnosis, informed consent waiver was granted by the IRB committee CMC, Vellore.

### Bacterial isolates, identification and antimicrobial susceptibility testing (AST)

Archived *S*. Enteritidis bloodstream infection (BSI) isolates (n = 101), collected between 2012 and 2022, were revived by culturing. Eight representative isolates from other clinical specimens (Feaces, Pus etc) were included in the study for the purpose of comparison. Isolates originating from poultry specimen was included to investigate potential zoonotic transmission. From poultry samples (rectal swabs and droppings), *Salmonella* sp. was selectively enriched by culturing in buffered peptone water and individual colonies were isolated using MSRV (Modified semi-solid Rappaport-Vassiliadis) media [18]. The isolates were further confirmed as *S*. Enteritidis by standard biochemical and sero-agglutination tests based on the Kauffmann-White scheme [19]. The isolates were further confirmed as *Salmonella* Enteritidis through seven-gene multilocus sequence typing (MLST) and comparison with the EnteroBase database (https://enterobase.warwick.ac.uk) [20].

AST (disk diffusion) was performed against ampicillin (10 μg), chloramphenicol (30 μg), co-trimoxazole (1.25/23.75 μg), ciprofloxacin (5 μg), and ceftriaxone (30 μg). Clinical breakpoints followed Clinical and Laboratory Standards Institute (CLSI) guidelines, 2022 [21]. Escherichia coli ATCC 25922 served as the quality control strain.

### DNA extraction and whole genome sequencing

A total of 48 clinical *S*. Enteritidis isolates were selected for WGS based on temporal distribution and phenotypic antimicrobial susceptibility profiles, while the remaining clinical isolates were excluded. Additionally, 17 poultry-derived isolates were included, resulting in a final set of 65 isolates for WGS. Genomic DNA was extracted using a Wizard DNA purification kit (Promega, Madison, USA) as per the manufacturer’s instructions. Paired-end fragment libraries were prepared using a Nextera XT DNA sample preparation kit following the manufacturer’s instructions (Illumina, Inc., San Diego, USA). The pooled libraries were diluted at a final concentration of 1.8 pM and sequenced on an Illumina Nextseq 500 platform with 150-cycle paired-end chemistry.

### Genome Assembly, Annotation and genotyping

Sequencing reads were evaluated with FastQC v0.12.0 (https://www.bioinformatics.babraham.ac.uk/projects/fastqc/) and reads with low sequencing quality were trimmed and removed using. Trimmomatic v0.39 (https://github.com/usadellab/Trimmomatic) was used to trim adapters and the internally corrected high-quality reads were subjected to genome assembly using SKESA v.2.4.0 (https://github.com/ncbi/SKESA). The assemblies were analysed for contamination using Kraken v2.1.3 (https://github.com/DerrickWood/kraken2) and quality was assessed via QUAST v5.2.0 (https://github.com/ablab/quast). Draft genome assemblies were examined using Seqsero v2.0 (https://github.com/denglab/SeqSero2) to confirm the antigenic profile of the serotype [22]. Sequence types were assigned *in silico* for all the isolates using the Multilocus sequence typing (MLST) pipeline available in the Center for Genomic Epidemiology (CGE) (https://cge.food.dtu.dk/services/MLST/). Antimicrobial resistance genes were screened using NCBI AMRFinderPlus v3.11.20 (https://github.com/ncbi/amr). Plasmids were identified from the genome data by searching against the PlasmidFinder database (https://cge.food.dtu.dk/services/PlasmidFinder/). The presence of pSEN-like plasmids and their associated genes was identified by performing BLAST comparisons against reference plasmids pSEN (HG970000) and pSENV (JN885080). Genome annotations were performed using Prokka v1.14.5 (https://github.com/tseemann/prokka) based on a custom genus database (https://github.com/tseemann/prokka?tab=readme-ov-file#crazy-person).

### Pangenome derived phylogeny

Annotated genomes (GFF3 files) generated by Prokka comprising study isolates (*n=65*) and a global representation of *S*. Enteritidis (*n=420*) were used as input to evaluate pan-genome diversity using Panaroo v1.3.4 (https://github.com/gtonkinhill/panaroo) [23]. Panaroo was run with standard parameters with ‘remove invalid genes’ enabled. Subsequently, single nucleotide polymorphisms (SNPs) were extracted from the core gene alignment generated by Panaroo using SNP-sites (http://sanger-pathogens.github.io/snp-sites/). A Maximum likelihood tree was constructed using IQTree2 (TVM+F+ASC+R2; bootstrap replicates = 100) [24] and visualized with iTOL (https://itol.embl.de/). Subsequently, distantly related outgroup isolates belonging to diverse sequence types were removed and a phylogenetic tree was reconstructed (*n=475*) to improve the cladistic structure. Phylogenetic clusters were assigned using rhierBAPS (https://github.com/gtonkinhill/rhierbaps) in its default mode (two cluster levels with 30 initial clusters). Minimum spanning trees (MSTs) were created and visualized using GrapeTree (https://achtman-lab.github.io/GrapeTree/MSTree_holder.html) from the core genome phylogeny [25]. Tree nodes were positioned through dynamic rendering and node style was adjusted by fine-tuning the node size and kurtosis. Nodes were coloured by the source of the isolates and node sizes were drawn proportionally to the number of isolates.

### Pangenome clustering

The gene presence or absence matrix created by Panaroo was clustered according to the four major phylogroups using the twilight analysis package (https://github.com/ghoresh11/twilight) using the default thresholds [26]. The pan genome reference FASTA file (pan_genome_reference.fa) generated by Panaroo, which encompasses all the genes within the dataset, was clustered using CD-HIT-EST. [27] with a 90% identity and 90% sequence length coverage. Finally, clustered phage-associated genes were predicted using PHASTEST database using default settings [28]. The prophage regions were further characterized through comparative analysis by aligning against reference sequences from the prophageDB database [29]. The predicted prophage regions were then manually curated to determine their gene composition and structural organization. Interactive visualization of the phylogenetic tree and PHASTER summary data was carried out in Phandango v1.3.0 [30].

### Lineage wise mutation profiling

Mutations were identified using *in-silico* analysis of single nucleotide polymorphisms (SNPs) using Snippy v4.6.0 mapping and variant calling pipeline (https://github.com/tseemann/snippy). To obtain the SNPs, we mapped the draft genomes (*n=475*) against the annotated features of the reference genome P125109 (NC_011294.1). We utilized custom-written bash scripts to extract the pattern of mutation accumulation with respect to the phylogenetic lineages. Genes that contained either frameshift mutation or a premature stop codon were manually curated and classified as hypothetically disrupted coding sequences (HDCS) or pseudogenes.

### Invasiveness index calculation

The potential of *S*. Enteritidis to cause invasive disease was calculated based on a machine learning pre trained invasive index predictive model (https://github.com/Gardner-BinfLab/invasive_salmonella) [31]. This approach classifies the genomes of invasive and gastrointestinal *Salmonella* by scoring deleterious mutations in 196 top predictor genes. The distribution of invasiveness index values for four major phylogroups was compared using the Kruskal-Wallis H Test.

### Statistical analysis

Statistical analysis was performed to investigate the potential association between severe immunosuppression and the clinical characteristics of *S*. Enteritidis infection. Data analysis was performed using GraphPad Prism 5.0 for Windows (GraphPad Software Inc). Continuous variables were represented as median ± interquartile range (IQR). Categorical variables were presented as either numbers or percentages. To compare these categorical variables, we employed either Fisher’s exact test or the Chi-squared test, as appropriate for the specific analysis. We calculated the odds ratio (OR) along with its corresponding 95% confidence intervals (CI). Statistical significance was determined by a P-value < 0.05 (two-tailed).

## Results

### Infants and immunosuppressed are more susceptible to *S*. Enteritidis BSI

The patient records indicated a total of 506 patients diagnosed with blood culture-positive iNTS disease between January 2012 and December 2022. Of these, 131 cases were confirmed to be caused by *S*. Enteritidis by conventional microbiological methods. After excluding 30 cases due to the absence of relevant clinical data and duplicate isolates (repeated samples from the same patient within 14 days), hospital records for 101 *S*. Enteritidis bacteraemia cases were analysed. The median age of affected patients was 34.5 years (IQR: 9–54.5 years) (Table 1). Notably, pediatric patients (<16 years) accounted for 27.72% (28/101) of cases, with infants (<12 months) comprising 13.86% (14/101), highlighting increased susceptibility in younger age groups. The most common presenting symptoms on admission included fever (75.25%), anemia/unhealthy appearance (59.41%), diarrhea (35.64%), and cough (30.69). Among 86 inpatients, the median fever duration was 6 days (IQR: 3–10 days). Male patients accounted for 65.34% (66/101) of cases.

We examined the impact of immunosuppressive conditions on *S*. Enteritidis bacteremia by comparing patients with these conditions to those without them. Among the 101 patients with *S*. Enteritidis bacteremia, 57.42% (58/101) had immunosuppressive conditions, including cancer/malignancy (24.75%), autoimmune disorders (21.78%), HIV infection (8.91%), and chronic steroid or chemotherapy use (39.6%) (Table 2). Among non-immunosuppressed patients (n=43), the pediatric population was disproportionately affected (44.18%), further emphasizing the vulnerability of children. Septic shock was significantly more frequent in immunosuppressed patients (27.59% vs. 9.3%; p = 0.025; OR = 3.71, 95% CI: 0.2 ± 0.078), indicating a higher risk of severe disease outcomes.

To evaluate treatment strategies and patient outcomes, we analyzed antibiotic therapy and recovery rates among affected individuals. A total of 65.34% (66/101) of patients received more than one antibiotic, primarily consisting of β-lactams (carbapenems, third-generation cephalosporins, β-lactam/β-lactamase inhibitors) and azithromycin. Clinical outcomes showed an overall recovery rate of 83.17% (84/101), while 10.89% (11/101) of patients died. Although mortality was higher in immunosuppressed patients (13.79%) compared to non-immunosuppressed patients (6.98%), this difference was not statistically significant (p = 0.345; OR = 2.13, 95% CI: 0.11 ± 0.0605).

### Phenotypic characterization

*S*. Enteritidis (*n=101*) strains were revived from the archived collections at the Department of Clinical Microbiology, CMC Vellore. Study isolates that were confirmed to be *S*. Enteritidis by biochemical testing and serotyping were subjected to AST. The results demonstrated that all isolates were susceptible to first-line antibiotics, (ampicillin, chloramphenicol, trimethoprim/sulfamethoxazole), and ceftriaxone. However, there was a decrease in susceptibility to fluoroquinolones, with 26.7% (27/101) of the isolates exhibiting non-susceptibility to ciprofloxacin.

### Population structure of *S*. Enteritidis consists of four major phylogenetic clades

The phylogenetic tree of 486 genomes was established by sequencing 65 isolates of *S*. Enteritidis (both clinical and poultry) and 421 publicly available genome sequences (**Fig. S1**). The metadata of isolates selected were described in **Table S1**. To improve branch length resolution, outgroup genomes belonging to diverse sequence types (ST180, ST1975, ST3304) were excluded, resulting in an abbreviated ML tree of 475 genomes. This dataset included 59 study isolates 42 from clinical sources and 17 from poultry along with genomes retrieved from public databases, allowing for a detailed assessment of the population structure of *S*. Enteritidis.

Phylogenetic analysis based on 13,876 core gene SNPs revealed that *S*. Enteritidis genomes clustered into four major phylogeneticclades (**Figure 1**). Clustering using RhierBAPS (Level 1) identified six BAPS clusters, four of which corresponded to the major phylogroups. In addition to previously defined groups namely the African lineage (BAPS Cluster 3), the global outlier/Atlantic lineage (Cluster 4), and the global epidemic clade (Cluster 1)—we identified a distinct phylogroup (BAPS Cluster 2), separated by 163 unique SNPs (**Table S2**). This novel cluster, which displays a geographically widespread global distribution, is hereafter referred to as the ‘Global Intermediate Clade’. Phylogenetically, this clade emerges as a paraphyletic group derived from the global outlier lineage, mirroring the evolutionary trajectory of the global epidemic clade. The study isolates were distributed between the Global Intermediate Clade (44.07%; 26/59), Global epidemic clade (28.8%; 17/59) and Global outlier clade (27.12%; 16/59). Notably, the majority of isolates were assigned to Sequence Type (ST)11, irrespective of their phylogenetic classification. Additionally, single-locus variants (SLVs) of ST11, including ST745, ST1479, and ST1974, were identified across the four primary clades.

**Figure 1:**
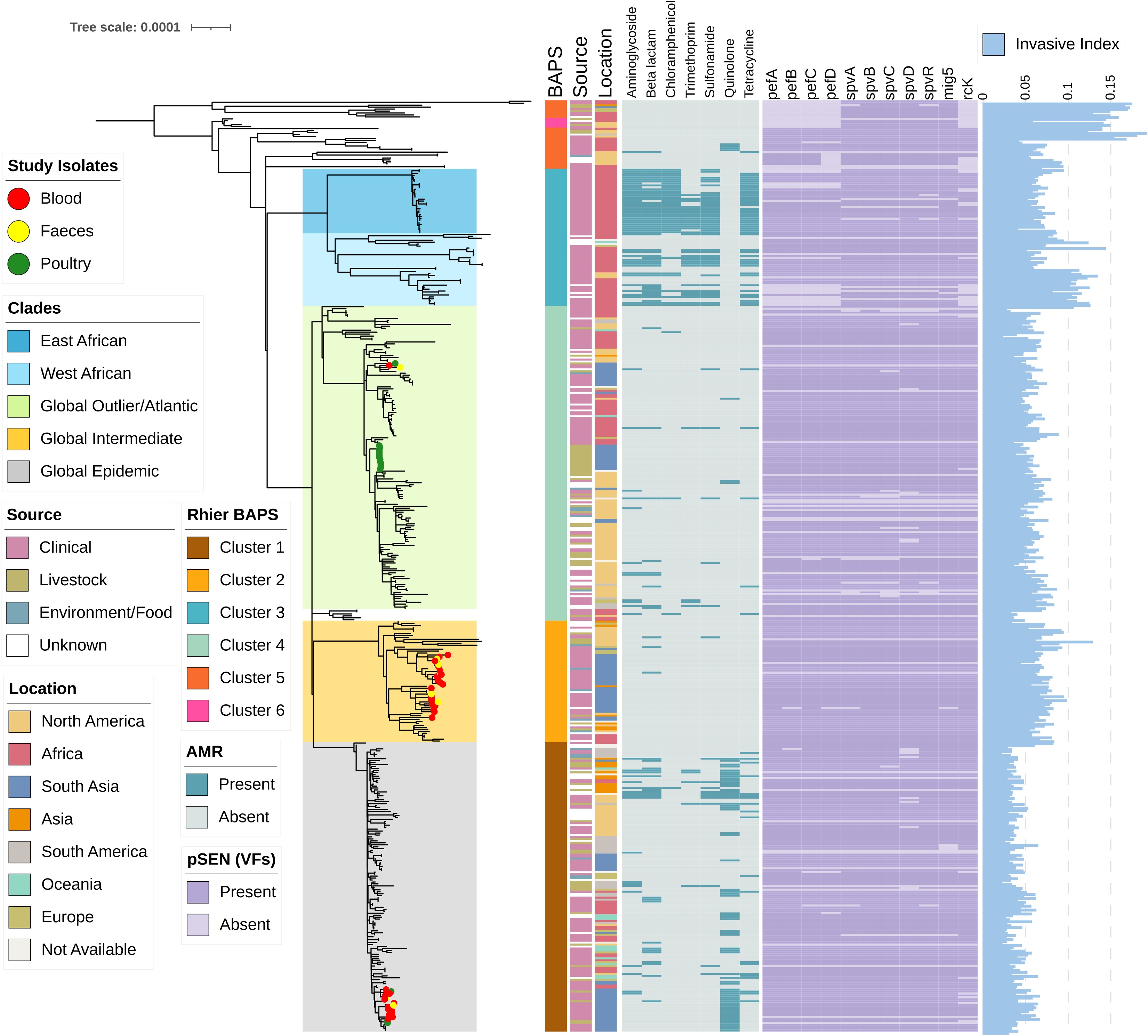
Maximum likelihood phylogenetic tree of *Salmonella* Enteritidis was constructed using 13,876 SNPs from 3,661 core genes (>95% presence) identified through pan-genome analysis with Panaroo (https://github.com/gtonkinhill/panaroo). The midpoint-rooted phylogeny was inferred using IQ-TREE2 (http://www.iqtree.org/), with bootstrap support values calculated from 100 replicates. The analysis included 475 *S*. Enteritidis genomes, of which 59 were isolates from this study. The phylogenetic tree reveals the distribution of *S*. Enteritidis within four main clusters, corresponding to distinct phylogroups. Study isolates are highlighted with red branch symbols. Metadata annotations are displayed as color strips: (1) BAPS clusters, (2) source of isolates, and (3) geographic location. Resistance profiles and key virulence genes are represented as heatmaps, while the invasiveness index for each genome is plotted as an external bar plot. The scale bar indicates substitutions per site. The tree was visualized and annotated using iTOL (https://itol.embl.de/).

The geographic distribution of *S*. Enteritidis phylogroups revealed distinct regional patterns. While some lineages exhibit global distribution, others appear geographically restricted. For instance, the African lineage includes West African and East African sub-lineages, both associated with iNTS disease. In contrast, the Global Outlier and Epidemic clusters contain isolates from diverse sources and geographic regions, spanning collections from 1925 to 2024 (**Table S3**). Notably, the Global Intermediate Clade appears to have originated in North America, with its earliest known isolate dating back to 1950 (SRR5819753). Ancestral strains of this lineage have been traced to the early 2000s, suggesting a subsequent expansion into Europe and Asia, reflecting its potential role in global *S*. Enteritidis dissemination.

### *S*. Enteritidis isolates from poultry were genetically distinct from those causing BSI

Comparative genome analysis was performed to investigate the genetic relationship between human and livestock isolates of *S*. Enteritidis and to assess potential zoonotic transmission pathways. Phylogenetic analysis revealed that 82.35% (14/17) of poultry isolates were assigned to the global outlier cluster while the remaining were grouped with the quinolone non-susceptible subcluster within global epidemic clade (**Figure S2)**. Among the poultry isolates clustered in the global outlier cluster, 13 formed a distinct subcluster that showed no direct genetic association with clinical isolates, suggesting limited direct transmission between poultry and human cases in this study. Notably, none of the poultry isolates of *S.* Enteritidis from the study area were found to be associated with the Global Intermediate clade (**Figure 1**).

### Limited antimicrobial resistance and plasmids among *S*. Enteritidis

Among the *S*. Enteritidis study isolates, we identified only a few genetic determinants of acquired antibiotic resistance. With the exception of one isolate, none of the study isolates contained detectable AMR genes, indicating a low prevalence of acquired resistance within this dataset. To provide a broader context, phylogroup-wise AMR screening was conducted on a global collection of isolates. Multidrug resistance (MDR) markers, including *bla*_TEM_, *cat*, *sul*, and *dfrA*, which confer resistance to first-line antibiotics, were primarily associated with East and West African clades. In contrast, AMR determinants showed variable prevalence across other phylogroups, regardless of their geographic origin. While the Global Epidemic Lineage occasionally carried genetic markers conferring resistance to ampicillin and co-trimoxazole, AMR genes were otherwise infrequent in non-African phylogroups. Unlike other NTS serovars, the majority of *S*. Enteritidis isolates remained susceptible to fluoroquinolones. However, quinolone resistance-conferring mutations in *gyrA* (S83Y or D87G) were identified in 25.42% (15/59) of study isolates, all of which belonged to the Global Epidemic Clade, suggesting the emergence of fluoroquinolone resistance within this lineage.

The pSEN-like virulence plasmid was commonly detected among global *S*. Enteritidis isolates. As expected, the pSENV plasmid and its associated replicon were identified in all isolates within the East African sub-clade. In contrast, a few of isolates from other phylogenetic clusters lacked the pSEN-like plasmid, though this absence appeared to be sporadic and not linked to phylogenetic lineage (**Figure S3**). Further analysis revealed the widespread presence of plasmid-encoded virulence factors (VFs) commonly associated with *S*. Enteritidis, including fimbrial adhesion genes (*pef*ABCD), type III secretion system (T3SS)-secreted effector genes, (*spv*BCD) encoded within *Salmonella* pathogenicity island 2 (SPI-2), and additional virulence genes (*mig-5* and *rck*) (**Figure 1**). Notably, the newly defined Global Intermediate Clade exhibited a high prevalence of these plasmid-associated virulence determinants, with 93.5% (58/62) of isolates carrying all eight genes, underscoring the potential role of this lineage in plasmid-mediated pathogenicity.

### African and Intermediate clade contained high levels of genome degradation

Frameshift mutations and premature stop codons leading to highly degraded coding sequences hypothetically disrupted coding sequences (HDCS) were analyzed across major phylogenetic clusters in the dataset (**Table S4**). Genomes within the African cluster, including both West African and East African sub-lineages, exhibited the highest number of HDCS (n=9) compared to isolates from the Global Epidemic and Global Outlier clusters (n=2). Additionally, representative genomes from the East African cluster contained an extra 16 HDCS, although none were associated with virulence genes. Isolates belonging to the Global intermediate clade were distinguished by eight lineage-defining HDCS (**Table S4**).

### Global intermediate clade has the second-highest invasiveness index, following the west African cluster

The invasiveness of *S.* Enteritidis phylogroups was evaluated using machine-learning based Invasive index calculator. This pipeline utilizes a random forest classifier model to produce predictive scores, denoted as delta bitscores (DBS). It accomplishes this by examining mutations within a previously defined set of 196 genes, as explained earlier. The potential of different phylogroups to cause extra-intestinal infection was compared using Wilcoxon– Mann–Whitney test. The results indicate that all examined *S.* Enteritidis phylogroups remain to be gastrointestinal (DBS <0.5). Nonetheless, the invasiveness index varies among these phylogroups (**Figure 2**). We observed a high invasiveness index (median = 0.253, SD = 0.25) for isolates belonging to West African clade while the global epidemic clade had the lowest (median = 0.188, SD = 0.01). The global intermediate clade also exhibited a relatively high invasiveness index (median 0.221, SD= 0.013) compared to East African and outlier clusters.

**Figure 2:**
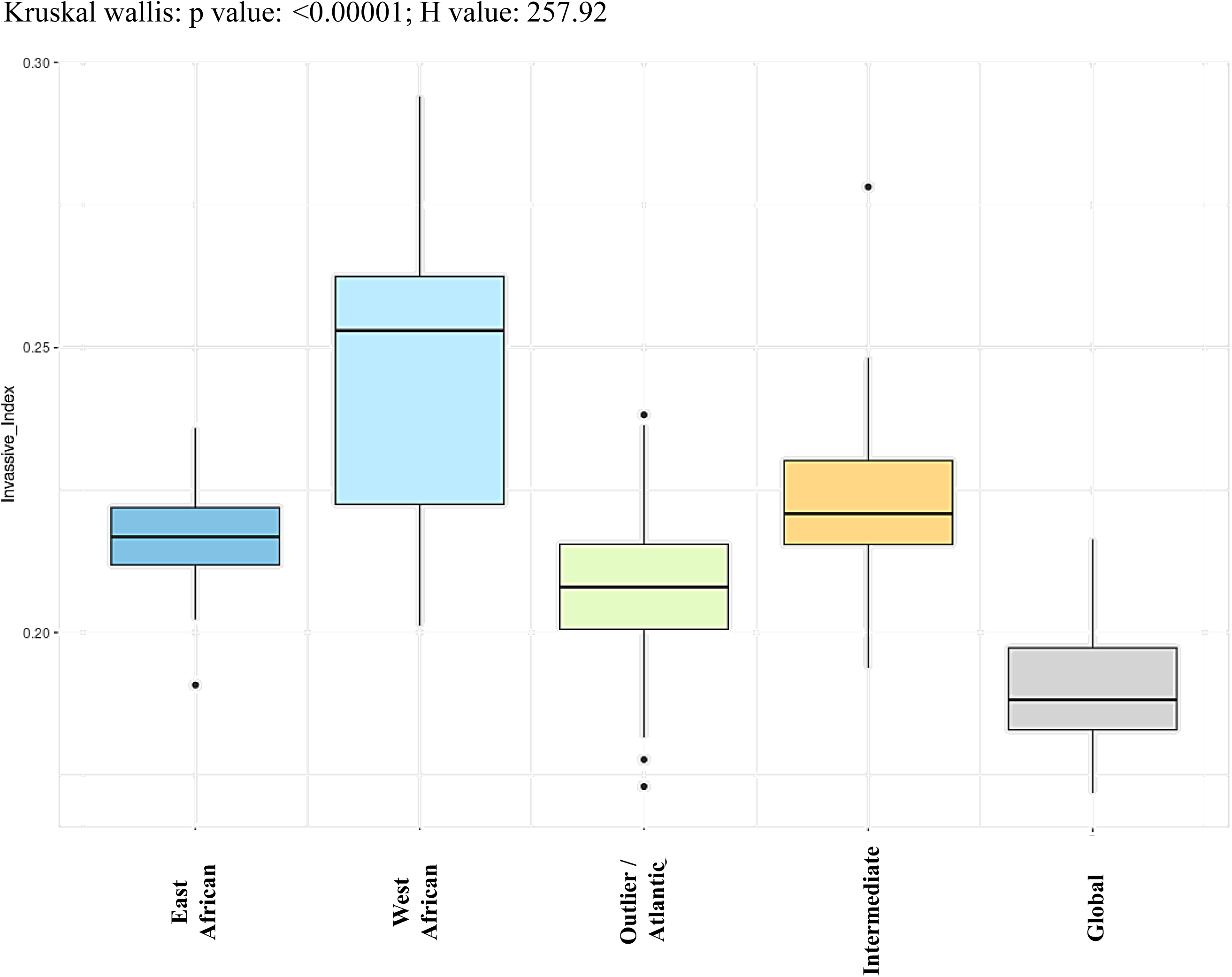
The lineage wise invasiveness index for all *Salmonella* Enteritidis genomes analyzed in this study was determined using the approach described by Wheeler et al. The median and standard deviation (SD) values for each lineage are as follows: East African (median = 0.217, SD = 0.010), West African (median = 0.253, SD = 0.026), Global Outlier (median = 0.208, SD = 0.011), Global Intermediate (median = 0.221, SD = 0.013), and Global Epidemic (median = 0.1883, SD = 0.010). Mann-Whitney U-test was used to compare differences between groups. In the boxplot, boxes represent the interquartile range (IQR), whiskers extend to 1.5× IQR, and outliers are shown as points. The Kruskal-Wallis test revealed significant differences across lineages (p < 0.0001, H = 257.92).

### Pan-genome clustering suggests the evolution of *S*. Enteritidis is largely driven by the acquisition of prophage genes

Pan-genome analysis of *Salmonella* Enteritidis, excluding highly divergent phylogroups (e.g., ST1975), identified 8,098 gene families. The core genome, comprising genes present in >99% of analyzed genomes, consisted of 3,672 genes, representing 73.8% of the total 5,344 genes. Analysis of the accessory genome revealed distinct gene family distribution patterns among established phylogroups (**Figure S4**).

Lineage-specific gene gain and loss events were particularly pronounced. The Global intermediate clade uniquely harbors a 56.9 kbp novel prophage (**Figure 3**), a mosaic structure exhibiting homology with Salmonella phage Sal3 (NC_031940) and Vibrio phage X29 (NC_024369). This prophage contains 62 coding sequences (CDS), including the virulence-associated *msgE* gene, suggesting a potential role in pathogenic adaptation. As reported previously, the African lineage has similarly acquired prophage, closely related to Enterobacter phage P88, in addition to the well-characterized Gifsy-1 prophage. In contrast, the emergence of the Global Epidemic Clade is associated with the acquisition of prophage ΦSE20, reinforcing the role of phage-mediated genetic changes in the evolution of epidemic *S*. Enteritidis strains.

**Figure 3:**
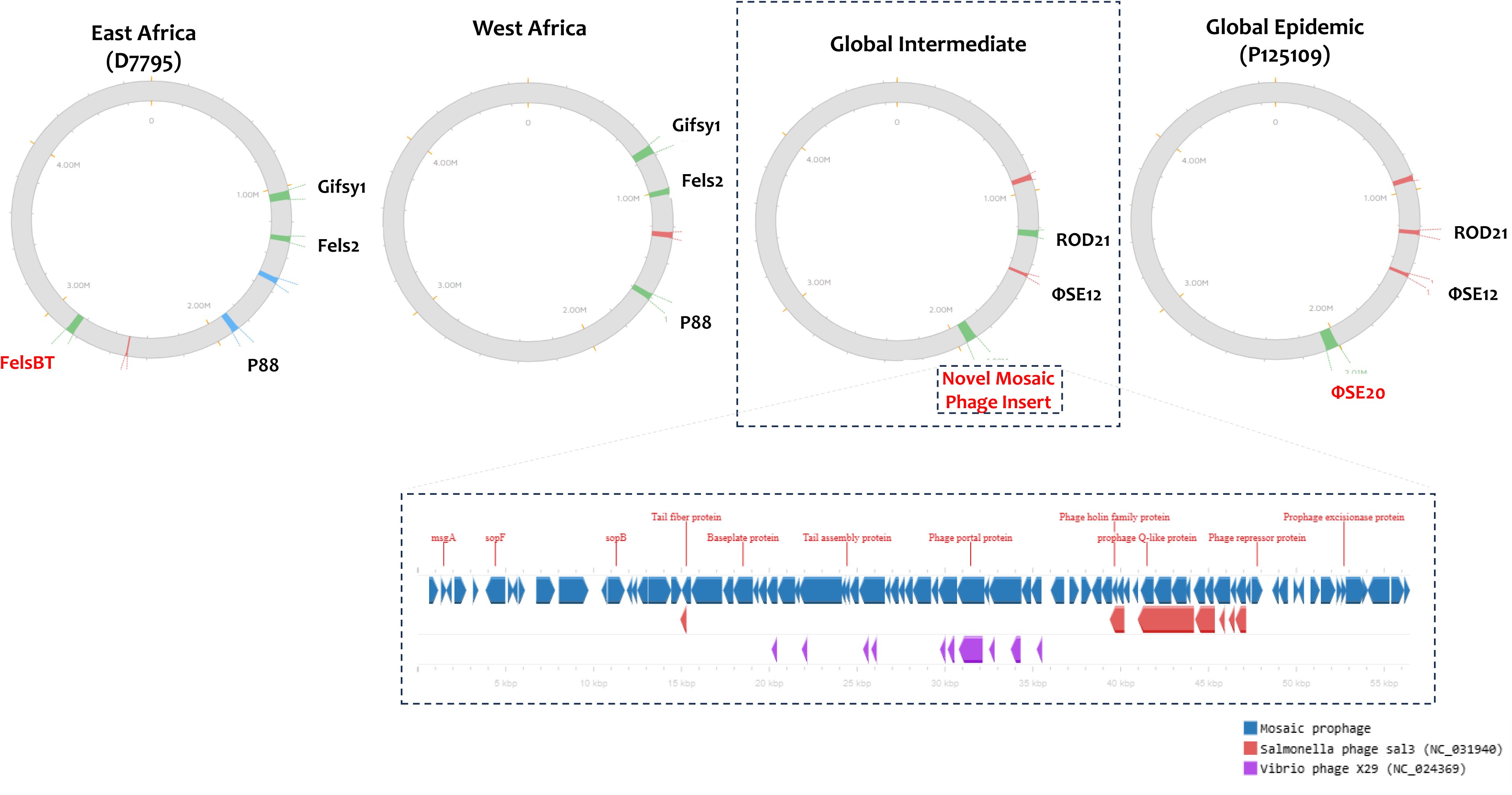
Prophage Map of Major *Salmonella* Enteritidis Lineages. The position of prophage regions identified in representative *S*. Enteritidis ST11 strains from each major lineage using PHASTER (https://phaster.ca/). The panel illustrates the genetic organization of a mosaic prophage identified in the global intermediate clade. Comparative genomic analysis of this mosaic prophage was performed using Proksee (https://proksee.ca/), showing its similarity to closest prophage homologs Salmonella phage Sal3 (NC_031940) and Vibrio phage X29 (NC_024369).

## Discussion

The increasing global incidence of iNTS infections, particularly BSI caused by *S*. Enteritidis, has been extensively documented in sSA [3, 6, 32, 33], However, its epidemiology in South Asia, including India, remains poorly characterized. In India, *S*. Enteritidis is the second most common iNTS serovar causing BSI, with prior studies also linking it to severe foodborne outbreaks (17, 34, 35]. This study presents the first comprehensive genomic and clinical analysis of *S*. Enteritidis isolates associated with BSIs in India, offering critical insights into their clinical features, phylogenetic relationships, and evolutionary mechanisms.

Many studies have identified immunosuppressive conditions such as HIV, malnutrition, and chronic illnesses as major risk factors for iNTS infections [14, 36, 37]. However, most of this evidence derives from African populations [6, 38]. Our study expands this understanding to the Indian context, demonstrating that infants and immunocompromised individuals (e.g., those with malignancies, autoimmune disorders, or undergoing invasive procedures) are at high risk of *S*. Enteritidis associated BSIs. Unlike in sub-Saharan Africa, where HIV coinfection drives iNTS [6], our cohort showed non-HIV-related immunosuppression as the dominant risk factor (Table 2). Pediatric cases comprised 27.7% of infections, with infants (<12 months) accounting for nearly half. Additionally, the high mortality rate (10.9%) and frequent empiric treatment with typhoid regimens (ceftriaxone-azithromycin) underscore the need for improved diagnostics and tailored management strategies for immunosuppressed and pediatric patients in India.

Recent genomic advancements have significantly improved our understanding of *S.* Enteritidis, elucidating its phylogenetic diversity, transmission dynamics, and mechanisms of virulence and antimicrobial resistance [13, 39–41]. Globally, sequence type (ST) 11 dominates, representing 90–94% of publicly available *S*. Enteritidis genomes [40], a trend reflected in our study, where the majority of isolates were assigned to ST11 (**Table S1**). Given the limited resolution of traditional 7-gene MLST classification, higher-resolution approaches such as core gene SNP based phylogenetics or core genome MLST (cgMLST) have enabled fine-scale differentiation of ST11 into distinct phylogenetic subclusters [13, 40]. Our phylogenetic analysis corroborates the presence of previously established global clades, including the regionally adapted East African and West African Clades (BAPS Cluster 5), the globally disseminated Outlier/Atlantic Clade (Cluster 2), and the Epidemic Clade (Cluster 4) (**Figure 1)** [13, 40, 42]. Interestingly, a substantial proportion of isolates from this study clustered within a previously undefined lineage (Cluster 1), which appears to have evolved from closely related progenitors within global lineages. EnteroBase metadata analysis (n=293) suggests that this lineage is widely distributed, with a significant proportion of isolates originating from Europe (n=109/293) and North America (n=80/293) (**Table S3**). Based on its unique phylogenetic placement and broad geographic distribution, we designate this lineage as the “Global Intermediate” Clade, with its earliest known isolate dating back to 1950s (SRR5819753).

Among the established *S*. Enteritidis clades the Central/East African and West African subclades exhibit distinct evolutionary trajectories, shaped by prophage acquisition, AMR determinants, and lineage-specific genomic degradation, all contributing to enhanced human host adaptation [13, 43]. For instance, the Central/East African Clade harbors 77 unique genes, including 33 plasmid-encoded (pSENV) virulence factors and 40 genes within a Fels-2-like prophage, while the West African clade contains 15 distinct genes, with 11 associated with a virulence plasmid. Notably, both African clades carry a ∼90 Kb virulence plasmid, nearly twice the size of the 58 Kb plasmid found in the Global Epidemic Clade, and encoding multiple AMR genes [44]. Similarly, the Global Epidemic Clade is distinguished by the presence of prophage φSE20, which is hypothesized to facilitate colonization in chickens, thereby enhancing transmission to humans and increasing disease potential [45]. These differences in prophage composition serve as key markers for distinguishing between major *S*. Enteritidis lineages [43, 46].

In contrast, the Global Intermediate Clade shares ROD21 and φSE12 prophages with the two other global lineages but also harbors a distinctive mosaic prophage (**Figure 3**). Genomic analysis of this novel prophage suggests that it may have diversified through horizontal gene transfer, acquiring genetic material from Salmon 118970 Sal3, Vibrio X29 and other prophages, while also maintaining unique genetic arrangement with no known similarity in current prophage databases. Given that this mosaic prophage is a defining feature of the Global Intermediate Clade, its emergence and diversification may have been driven majorly by phage-associated gene flux [47].

The genetic basis of invasive *Salmonella* lineages remains incompletely understood; however functional diversification through genome degradation and acquisition of virulence factors appears to be key drivers of host adaptation [48]. In our study, the Global intermediate clade displayed an elevated invasiveness index (DBS), closer to the two African Clades (**Figure 2)**, suggesting that these lineages may have undergone adaptive genomic changes favoring extraintestinal survival and infection. Since loss-of-function mutations and the accumulation of HDCS are hallmarks of host-adapted Salmonella lineages [49] the increased invasive index divergence in the intermediate clade, coupled with evidence of genomic degradation, supports its potential adaptation to infect humans. These findings, alongside epidemiological landscape of ST313 *S*. Typhimurium, strongly suggest that the *S.* Enteritidis Global intermediate clade has diversified from global epidemic clade for enhanced human infectivity [12].

The global spread of *S*. Enteritidis is closely linked to contaminated poultry products, which played a key role in its introduction and worldwide dissemination (42, 46]. Genome-based source tracking suggests a centralized origin, likely emerging during the early industrialization of poultry production [16, 50]. Comparative genomic analysis of *S*. Enteritidis isolates from clinical and poultry sources in India reveals that poultry-derived isolates cluster within the Outlier/Atlantic and Global Epidemic Clades, aligning with previous reports linking these lineages to poultry-driven transmission [42]. Notably, no contemporary poultry isolates are associated with the Global Intermediate Clade, which predominantly comprises Indian bloodstream infection (BSI) isolates. This pattern mirrors the African epidemic lineages, where invasive human-adapted strains evolved independently of poultry-associated strains. Thus, the Global Intermediate Clade appears to be primarily linked to human infections, while poultry isolates cluster within region-specific subclades of the Outlier and Global Epidemic lineages.

Our study has several limitations. First, clinical data originate from a single Indian hospital, limiting generalizability to the national disease burden. Second, most clinical variables lacked statistical association with infection outcomes. Third, phylogenetic analysis included 65 of 101 isolates from the study period, with underrepresentation of poultry-derived isolates restricting epidemiological inferences. Finally, functional characterization of the identified prophages, though beyond this study’s scope, is needed to confirm their evolutionary significance. Despite these limitations, our data provide a robust evaluation of global phylogenomics for this understudied pathogen.

### Conclusion

Our findings align with current knowledge regarding the clinical presentation, evolutionary processes, and transmission patterns of *S*. Enteritidis. Severely immunosuppressed individuals are at higher risk of BSI caused by NTS serovars, reinforcing the opportunistic nature of these infections. Our phylogenomic analyses reveal that Indian invasive *S*. Enteritidis isolates mostly belong to an emerging lineage, which we designate as the Global Intermediate Clade. This clade exhibited high invasiveness, second only to the West African Clade, and was characterized by a distinct prophage repertoire and extensive genomic degradation. Given its genetic divergence from poultry-associated strains, these findings suggest that this lineage may have undergone adaptation to cause bloodstream infections in humans. Further research is needed to better understand the environmental reservoirs and transmission pathways of the Global Intermediate Clade.

## Data Availability

Raw read data were deposited in the European Nucleotide Archive (ENA) under project accession number: PRJEB87860. The individual sample accession numbers are listed in Table S1

https://www.ncbi.nlm.nih.gov/bioproject/PRJEB87860/

## Acknowledgements

We express our heartfelt gratitude to the Department of Clinical Microbiology, Christian Medical College, Vellore, for providing the essential facilities and support that made this study possible. We extend our sincere appreciation to the clinicians, Dr. Jithin Joy and Dr. Suriya Chandran, for their efforts in contributing to the clinical data collection. We are also grateful to Ms. Baby Abirami Shankar and Mr. Ayyanraj N. for their valuable assistance with phenotypic testing, stock culture maintenance, and sequencing. Special thanks go to Ms. Agila K. Pragasam for her expert guidance and support in the genomic analysis.

## Ethical Clearance

Institutional Review Board (IRB) of Christian Medical College (CMC), Vellore, India gave ethical approval for this work vide IRB Min no. 12626 dated 26.02.2020. As the required data had been collected as part of the standard of care for diagnosis, informed consent waiver was granted by the IRB committee CMC, Vellore.

## Contributors

Conceptualization: JJJ, AV, BV and KW

Methodology and Investigation: JJJ, AV, DS, RI, KG, JRM, YM, VM and BLY

Data analysis: JJJ and AV

Visualization: JJJ and AV

Funding acquisition: BV and KW

Project administration: JJJ, DS, YM, FA, MPT, SK

Supervision: DS, BV and KW

Original draft: JJJ, AV

Review & editing: JJ, BV and KW

## Disclaimer

The funders had no role in the design and conduct of the study; collection, management, analysis, and interpretation of the data; preparation, review, or approval of the manuscript; and decision to submit the manuscript for publication.

## Patient consent for publication

Not required.

---

**Figure S1:** Pan-genome based core genome phylogenetic tree of 486 *S*. Enteritidis showing the comparative phylogenetic clustering by MLST. Study isolates are highlighted with red branch symbols. MLST are displayed as color strips. The tree was visualized and labeled using iTOL (https://itol.embl.de/).

**Figure S2:** Minimum spanning tree (MST) of *S*. Enteritidis isolates (*n=475*) constructed and visualized using GrapeTree (https://achtman-lab.github.io/GrapeTree/MSTree_holder.html). Each node represents a unique ST or Clonal group, and node size is proportional to the number of isolates with that ST. Node color indicates the source from which the isolate was obtained. Tree nodes were positioned through dynamic rendering and node style was adjusted by fine-tuning the node size and kurtosis.

**Figure S3:** Phandango heatmap illustration (Hadfield et al. 2018) focusing on the presence of AMR genes, virulence genes and pSEN-like plasmid associated genes across phylogenetic lineages. The orange indicates the presence of a gene and purple indicates its absence.

**Figure S4:** Pangenome analysis of 475 *S*. Enteritidis isolates indicates the diversity in accessory genomes among the different phylogenetic lineages. Phandango heatmap illustration (Hadfield et al. 2018) of gene presence and absence matrix (right side) against Maximum likelihood phylogenetic tree (Left side) with clades correlate with the observation derived from phylogenetic analysis. The orange indicates the presence of a gene and purple indicates its absence. A highlighted region (white dashed rectangle) indicates a cluster of lineage-specific genes.

