## Supplementary figures and images for "Pan genome clustering identifies a novel mosaic prophage specific to *Salmonella* Enteritidis lineage associated with the invasive disease in India"

### Fig.S1, Fig.S2, Fig.S3, Fig.S4

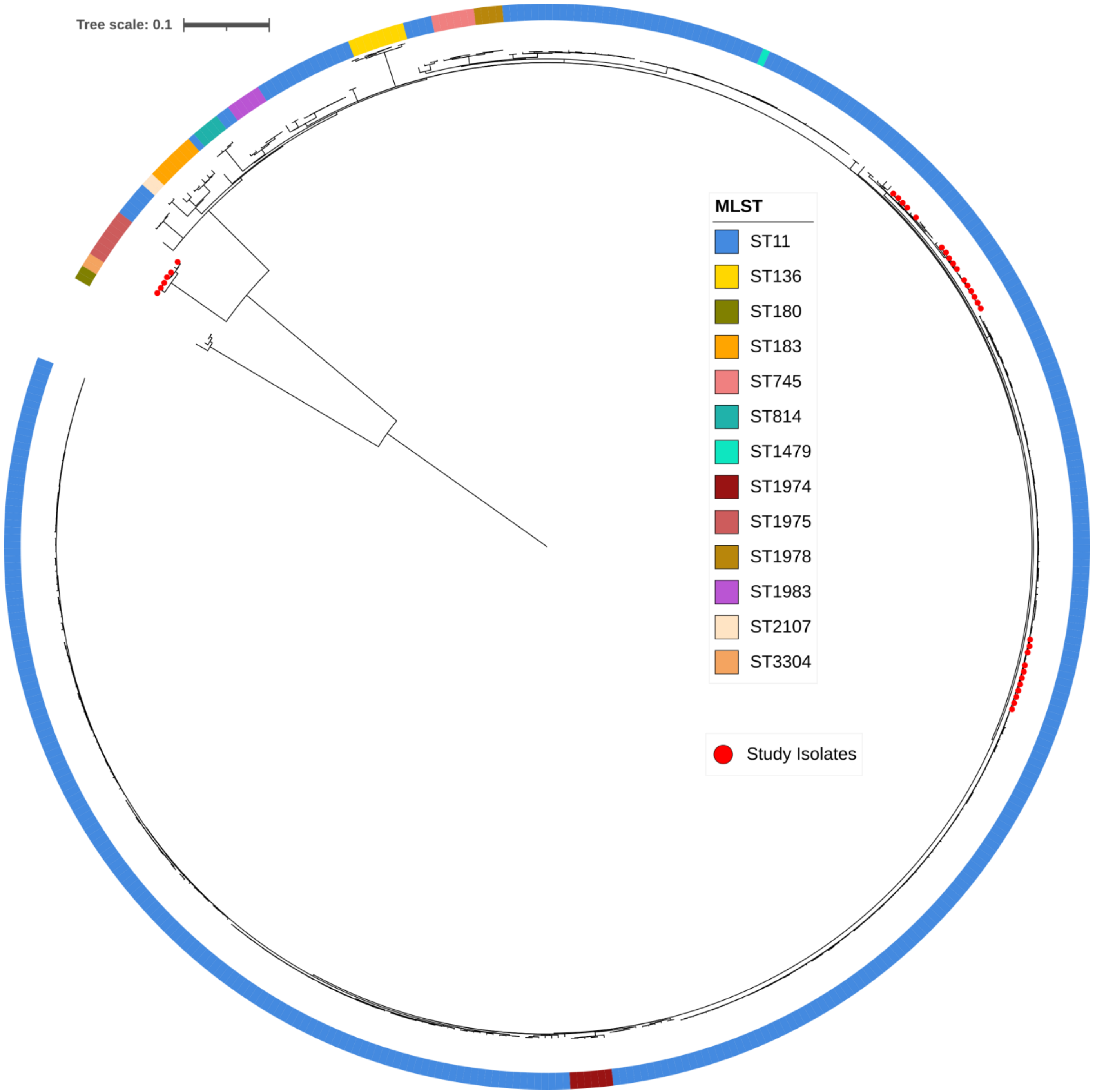

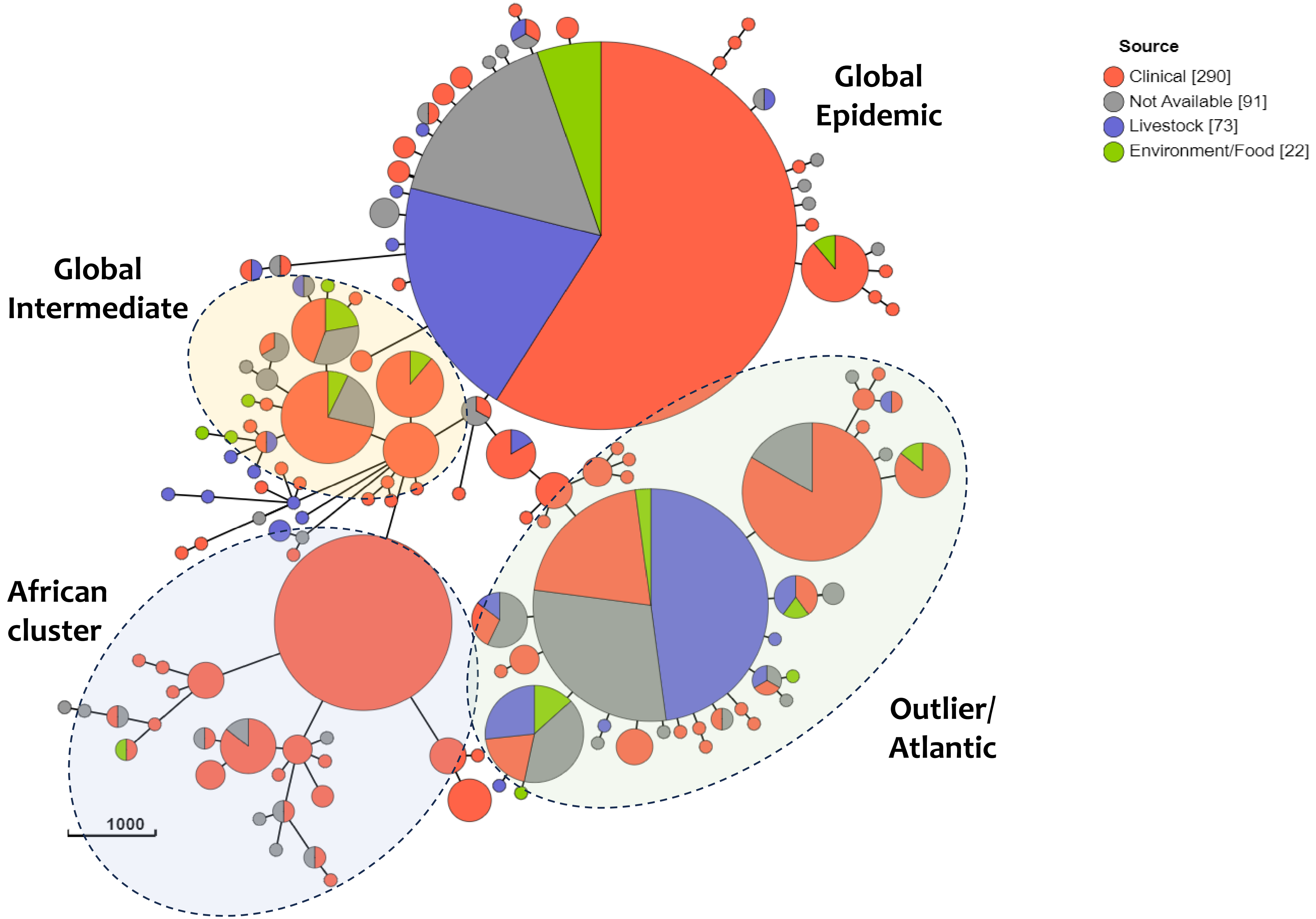

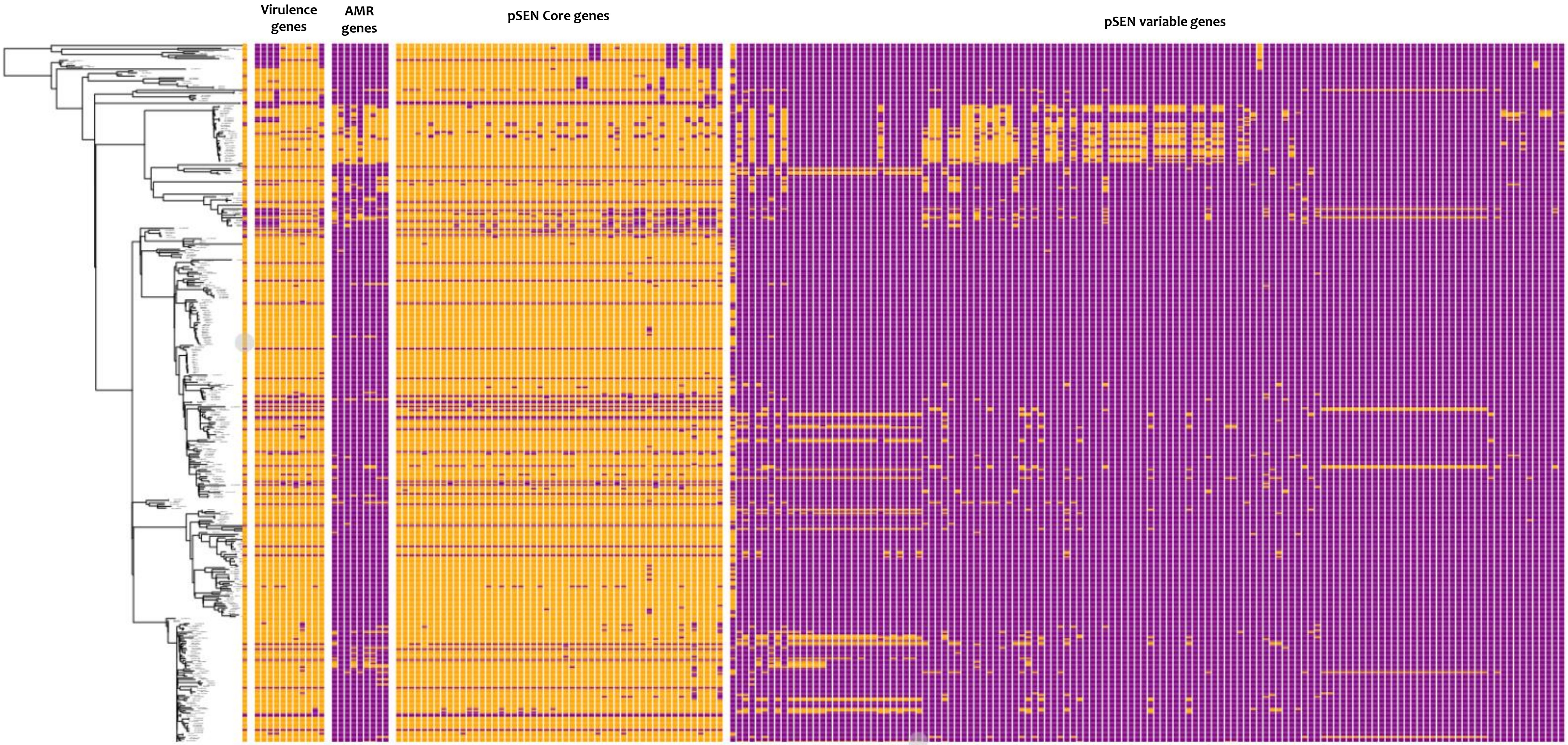

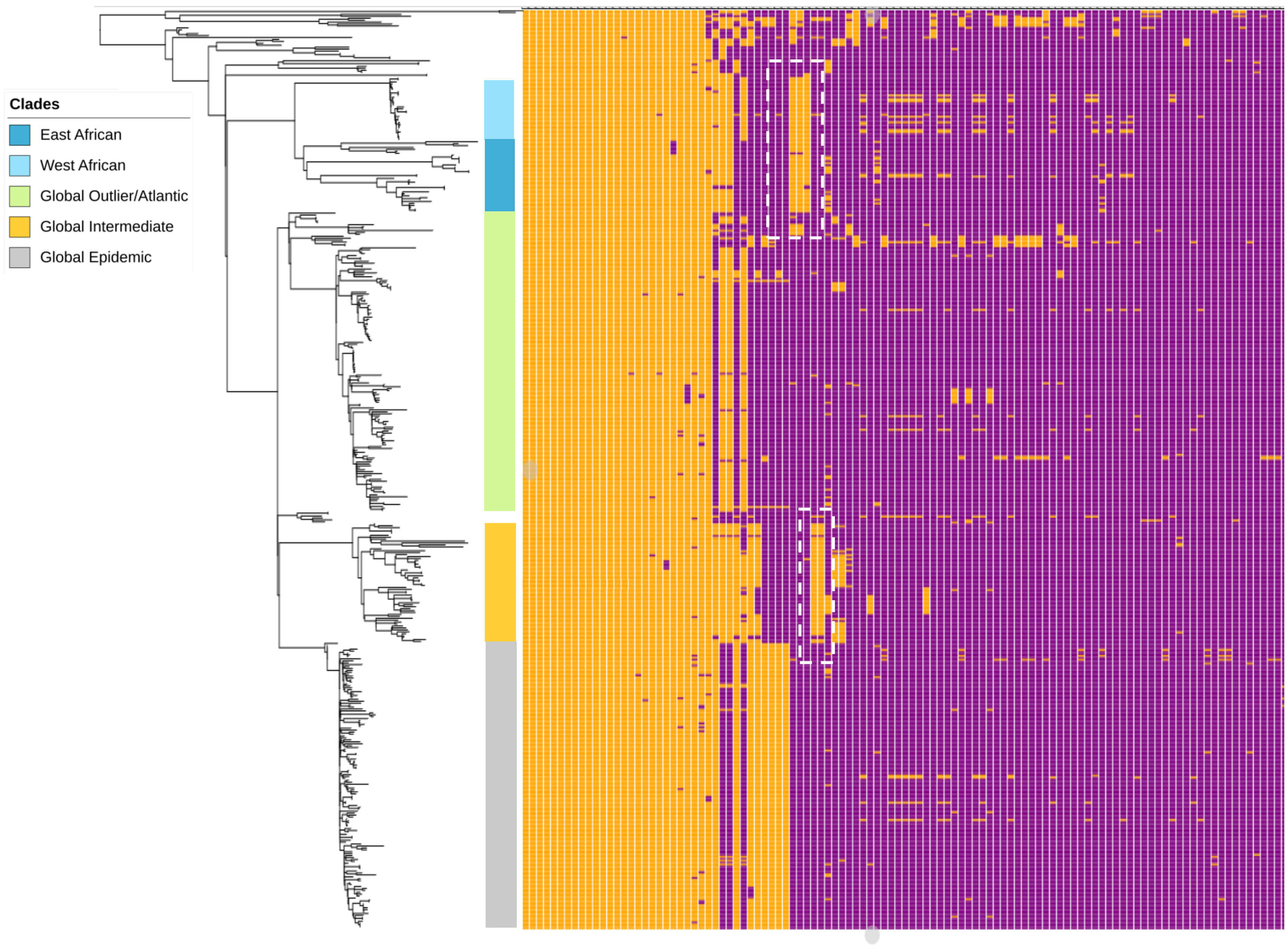
